# Incidence and etiology of clinically-attended, antibiotic-treated diarrhea among children under five years of age in low- and middle-income countries: evidence from the Global Enteric Multicenter Study

**DOI:** 10.1101/2020.05.11.20098640

**Authors:** Joseph A. Lewnard, Elizabeth T. Rogawski McQuade, James A. Platts-Mills, Karen L. Kotloff, Ramanan Laxminarayan

**Affiliations:** Division of Epidemiology, School of Public Health, University of California, Berkeley, Berkeley, California, United States; Division of Infectious Diseases and Vaccinology, School of Public Health, University of California, Berkeley, Berkeley, California, United States; Center for Computational Biology, College of Engineering, University of California, Berkeley, Berkeley, California, United States; Department of Public Health Sciences, University of Virginia School of Medicine, Charlottesville, Virginia, United States; Division of Infectious Diseases and International Health, University of Virginia School of Medicine, Charlottesville, Virginia, United States; Department of Pediatrics, Center for Vaccine Development and Global Health, University of Maryland School of Medicine, Baltimore, Maryland, United States; Center for Disease Dynamics, Economics & Policy, New Delhi, India; Princeton Environmental Institute, Princeton University, Princeton, New Jersey, United States

## Abstract

**Background:** Diarrhea is a leading cause of antibiotic consumption among children in low- and middle-income countries. While vaccines may prevent diarrhea infections for which children often receive antibiotics, the contribution of individual enteropathogens to antibiotic use is minimally understood. We used data from the Global Enteric Multicenter Study (GEMS) to estimate pathogen-specific incidence of antibiotic-treated diarrhea among children under five years old residing in six countries of sub-Saharan Africa and South Asia before rotavirus vaccine implementation.

**Methods and findings:** GEMS was an age-stratified, individually-matched case-control study. Stool specimens were obtained from children presenting to sentinel health clinics with newly-onset, acute diarrhea (including moderate-to-severe and less-severe diarrhea) as well as matched community controls without diarrhea. We used data from conventional and quantitative molecular diagnostic assays applied to stool specimens to estimate the proportion of antibiotic-treated diarrhea cases attributable to each pathogen. Antibiotics were administered or prescribed to 9,606 of 12,109 moderate-to-severe cases and 1,844 of 3,174 less-severe cases. Across all sites, incidence rates of clinically-attended, antibiotic-treated diarrhea were 12.2 (95% confidence interval: 9.0-17.8), 10.2 (7.4-13.9) and 1.9 (1.3-3.0) episodes per 100 child-years at risk at ages 6 weeks to 11 months, 12-23 months, and 24-59 months, respectively. Based on the recommendation for antibiotic treatment to be reserved for cases with dysentery, we estimated a ratio of 12.6 (8.6-20.8) inappropriately-treated diarrhea cases for each appropriately-treated case. Rotavirus, adenovirus serotypes 40/41, *Shigella*, sapovirus, Shiga toxin-producing *Escherichia coli*, and *Cryptosporidium* were the leading antibiotic-treated diarrhea etiologies. Rotavirus caused 29.2% (24.5-35.2%) of antibiotic-treated cases, including the largest share in both the first and second years of life. *Shigella* caused 14.9% (11.4-18.9%) of antibiotic-treated cases, and was the leading etiology at ages 24-59 months.

**Conclusions:** Our findings should inform the prioritization of vaccines with the greatest potential to reduce antibiotic exposure among children.

**AUTHOR SUMMARY:** Because diarrhea is the second-leading cause of antibiotic consumption among children in low- and middle-income countries (LMICs), effective vaccines against diarrheal pathogens may have the collateral benefit of reducing antibiotic exposure and resistance selection in these settings. Whereas antibiotic treatment is only recommended for diarrhea cases with blood in the stool (which suggests *Shigella* etiology), little is known about real-world diarrhea treatment practices in LMICs. We used data from a study of children experiencing diarrhea and matched, asymptomatic controls residing in LMICs of South Asia and sub-Saharan Africa to understand factors leading to antibiotic treatment of diarrhea cases, and the proportion of cases attributable to various enteric pathogens. We identify rotavirus and *Shigella* as the predominant causes of antibiotic-treated diarrhea at ages 0-23 months and 24-59 months; additional leading causes include adenovirus serotypes 40/41, sapovirus, Shiga toxin-producing *Escherichia coli*, and *Cryptosporidium*. We estimate that approximately 12.6 diarrhea cases receive antibiotics inappropriately for each appropriately-treated case in the study settings. Vaccines against prominent enteropathogens may substantially reduce antibiotic consumption among children in LMICs.

## INTRODUCTION

Prevention of avoidable antibiotic consumption is a central goal of efforts to counter the growing threat of antimicrobial resistance [1]. Childhood vaccines, which have been successful in preventing deaths among children under five years of age globally [2], may have an important role to play in achieving this goal. Acute respiratory infections and diarrhea are the leading causes of antibiotic consumption among children in low- and middle-income countries (LMICs) [3]. Little is known about factors that lead to antibiotic treatment of acute disease cases in these settings, and the extent of antibiotic use that is appropriate or inappropriate with respect to disease etiology [4,5]. These uncertainties pose a challenge to quantifying vaccine-preventable antibiotic consumption as a component of the impact of vaccines on antimicrobial resistance [6,7].

Randomized and observational studies in high-income settings have reported that vaccines against influenza [8] and *Streptococcus pneumoniae* [9–11] reduce antibiotic use by preventing acute respiratory infections. Despite prevalent antibiotic treatment of diarrhea, especially in LMICs [3], there have been no similar assessments of the impact that enteropathogen vaccines may achieve against antibiotic consumption [12]. Evidence of the pathogen-specific burden of antibiotic-treated diarrhea is needed to prioritize investments in development and implementation of vaccines with the greatest potential impact.

The Global Enteric Multicenter Study (GEMS) [13–16] was undertaken before rotavirus vaccine implementation in seven LMICs across sub-Saharan Africa and South Asia to determine the burden and etiology of diarrhea among children under five years old in these settings. Clinical data from the study provide a view of the management of diarrhea cases in LMIC healthcare settings, including antibiotic treatment practices. We revisited data from GEMS aiming to characterize real-world diarrhea treatment practices in LMICs, and to inform vaccine prioritization by estimating pathogen-specific incidence of clinically-attended, antibiotic-treated diarrhea.

## METHODS

### Study design

GEMS was a multi-site, age-stratified, individually-matched case-control study characterizing the incidence, etiology, and clinical consequences of diarrhea among children at four sites in sub-Saharan Africa (Basse, The Gambia; Bamako, Mali; Manhiça, Mozambique; and Siaya County, Kenya) and three sites in South Asia (Mirzapur, Bangladesh; Kolkata, India; and Bin Qasim Town, Karachi, Pakistan. Participants were children ages 6 weeks to 59 months who belonged to routinely-censused populations, with demographic surveillance systems in place to record all births, deaths, and migrations. Enrollment of cases occurred at sentinel hospitals and health centers where children sought care for diarrhea. The design, procedures, and primary outcomes have been reported previously [13–15,17]. Briefly, all children seeking care at sentinel health centers were screened for diarrhea, defined as ≥3 loose stools in the preceding 24 hours; cases were considered eligible if newly-onset (within the preceding 7 days) and acute (preceded by ≥7 diarrhea-free days) illness was reported. Moderate-to-severe diarrhea (MSD) cases met at least one of the following criteria, as assessed by study clinicians: sunken eyes (confirmed by the parent or caretaker as unusual); decreased skin turgor (abdominal skin pinch with slow or very slow [>2s] recoil); intravenous hydration administered or prescribed; visible blood in loose stools (dysentery) reported by the parent, clinician, or laboratory; or hospitalization. Acute, new-onset diarrhea cases not meeting the above criteria were considered to be less-severe diarrhea (LSD).

The study included a 3-year primary phase (GEMS-1) enrolling MSD cases as well as a 1-year follow-on phase (GEMS-1A) enrolling both MSD and LSD cases; due to an anticipated low enrollment, LSD cases were not enrolled in the Kenya site. Study personnel aimed to enroll the first 9 children with MSD (in the 3-year primary study) and the first 9 children with MSD or LSD (in the 1-year follow-on study) at each site, over each fortnight, within each of three age strata: infants ages 6 weeks to 11 months, toddlers ages 12-23 months, and young children ages 24-59 months. For each case, investigators aimed to enroll 1-3 matched controls from the community. Controls of the same age (±2 months for cases ages ≤23 months and ±4 months for cases ages 24-59 months), sex, and village or neighborhood were selected at random from demographic surveillance site databases and enrolled within 14 days of the case at home visits by field workers. Controls experiencing diarrhea in the preceding 7 days were ineligible. Standardized demographic, epidemiologic, and clinical information was collected by study personnel at enrollment, in consultation with parents or primary caretakers of children.

We defined clinically-attended, antibiotic-treated diarrhea cases as MSD or LSD cases who were administered antibiotics during the enrollment visit, or who received an antibiotic prescription at the enrollment visit. From this definition, we excluded cases diagnosed with other conditions that would justify antibiotic treatment, regardless of diarrhea symptoms; we considered these diagnoses to include pneumonia/lower respiratory-tract infection, meningitis or other invasive bacterial infection, and typhoid.

### Enteropathogen assessment

Microbiological procedures in GEMS have been described previously [18]. Briefly, ≥3 grams of fresh stool was collected from each participant and placed in cold storage within 1 hour; additionally, 2 rectal swabs were obtained for bacterial cultures from children who were going to receive antibiotics before stool passage. Specimens were placed into transport media and processed within 18 hours. Putative enteropathogens included *Salmonella* spp., *Shigella* spp., *Campylobacter* spp., *Aeromonas* spp., *Vibrio cholerae*, diarrheogenic *Escherichia coli*, rotavirus, adenovirus (serotypes 40 and 41), norovirus (serotypes GI and GII), sapovirus, astrovirus, *Giardia lamblia, Entamoeaba histolytica*, and *Cryptosporidium* spp.

For this study, we used data obtained from both conventional diagnostic procedures (available from all children) and quantitative molecular diagnostic assays (available from a subset of 5034 sampled pairs of participants in the primary study, as detailed below) [16]. Under conventional diagnostic approaches, bacterial pathogens were detected using standard culture techniques. A multiplex polymerase chain reaction (PCR) assay was used to type *E. coli* isolates as enteropathogenic, enteroaggregative, and Shiga toxin-encoding enterotoxigenic (ST-ETEC; defined as encoding *eltB* for heat-labile toxin, *estA* for heat-stable toxin, or both *eltB* and *eltA*). Rotavirus, adenovirus, *G. lamblia, E. histolytica*, and *Cryptosporidium* spp. were detected via commercial immunoassays, and norovirus, sapovirus, and astrovirus were detected via multiplex reverse transcriptase (PCR).

Stool specimens from up to 300 randomly-selected MSD cases and the first available matched controls for each age stratum and study site from the primary study were retested using a custom TaqMan Array Card (Thermo Fisher, Carlsbad, USA) with probe-based quantitative PCR (qPCR) assays for 32 enteropathogens, as described previously [16,19]. The number of quantification cycles (Cqs) at which fluorescence exceeded background rates was recorded for each pathogen. Detections with Cq≥35 were considered to be negative.

We estimated adjusted attributable fractions using models that included the Cq value for each pathogen as covariates, similar to previous analyses [16,20]; we detail statistical approaches in the supplementary methods (**S1 Text**). Analyses were stratified for MSD and LSD cases. We also estimated adjusted attributable fractions using pathogen detection data from conventional assays, as applied in the primary analyses of the GEMS datasets [13,14,21]. Dividing adjusted attributable fraction estimates based on qPCR data for clinically-attended, antibiotic-treated MSD cases, by estimates generated from conventional diagnostic data among such cases, yielded a multiplier representing the relative burden that we would expect to estimate if qPCR assays had been undertaken for all cases. We used these multipliers to rescale adjusted attributable fraction estimates for clinically-attended antibiotic-treated LSD cases, for whom only conventional diagnostic data were available.

### Healthcare utilization and attitudes surveys

An initial cross-sectional healthcare utilization and attitudes survey was conducted within random samples of children belonging to the demographic surveillance population of each site, followed by brief serial surveys to document in-migration, out-migration, births, and deaths, as described previously [17]. Parents or caretakers were interviewed to determine whether children had experienced acute, newly-onset MSD or LSD within 7 days preceding the interview, and whether care was sought care at a sentinel hospital or health center within 7 days of diarrhea onset. We used these data to estimate incidence of diarrhea within the population of each study site, following the original analysis protocol [21] (**S1 Text**).

### Additional analyses

Because antibiotic treatment of dysentery is recommended based on the possibility of *Shigella* etiology, we also estimated adjusted attributable fractions of clinically-attended, antibiotic-treated diarrhea associated with dysenteric and non-dysenteric *Shigella* infections. In addition, we estimated the ratio of incidence of inappropriate (i.e., non-dysenteric) to appropriate (dysenteric) antibiotic-treated cases in each setting. To understand differences in symptoms and management of diarrheal infections, we further estimated associations of detection of each pathogen with administration or prescription of an antibiotic (among MSD and LSD cases) and with dysentery and hospitalization (among MSD cases only). We detail the statistical approaches for these analyses in the supporting information (**S1 Text**).

### Ethics

The clinical protocol of the original study was approved by ethics committees of the University of Maryland, Baltimore, Maryland, United States, and at each of the participating field sites; parents or primary caretakers provided written informed consent for participating children at enrollment. This secondary analysis of the de-identified GEMS dataset was considered to be non-human subjects research and was therefore exempt from full review by the Committee for Protection of Human Subjects of the University of California, Berkeley.

## RESULTS

### Enrollment and antibiotic treatment of diarrhea cases

Data were available from 12,109 cases with MSD seeking care at sentinel hospitals and health centers recruited in the primary and follow-on studies between December, 2007 and November, 2012 [13]. The one-year follow-on study recruited 3,174 LSD cases across all sites except Siaya County, Kenya.

In total, 9,606 children with MSD and 1,844 children with LSD were administered or prescribed antibiotics (**Table 1**). Concordant with World Health Organization guidelines [22,23] for antibiotic treatment of dysentery, 95.6% (2,345/2,454) of dysenteric cases were administered or prescribed antibiotics. In Bangladesh, where roughly half (1,283/2,454) of all dysentery cases in the study occurred, 99.5% of cases (1,276/1,283) were administered or prescribed antibiotics. In total, 1.8% (43/2,454) of cases experiencing dysentery were diagnosed with other conditions justifying antibiotic treatment, versus 8.8% (852/9,655) of cases with non-dysenteric MSD and 2.2% (69/3,174) of LSD cases (**S2 Table**). Contrary to current guidelines [22,23], 73.9% (6,505/8,803) of non-dysenteric MSD cases and 57.7% (1,791/3,105) of LSD cases were administered or prescribed antibiotics in the absence of other diagnoses justifying such treatment.

**Table 1:**
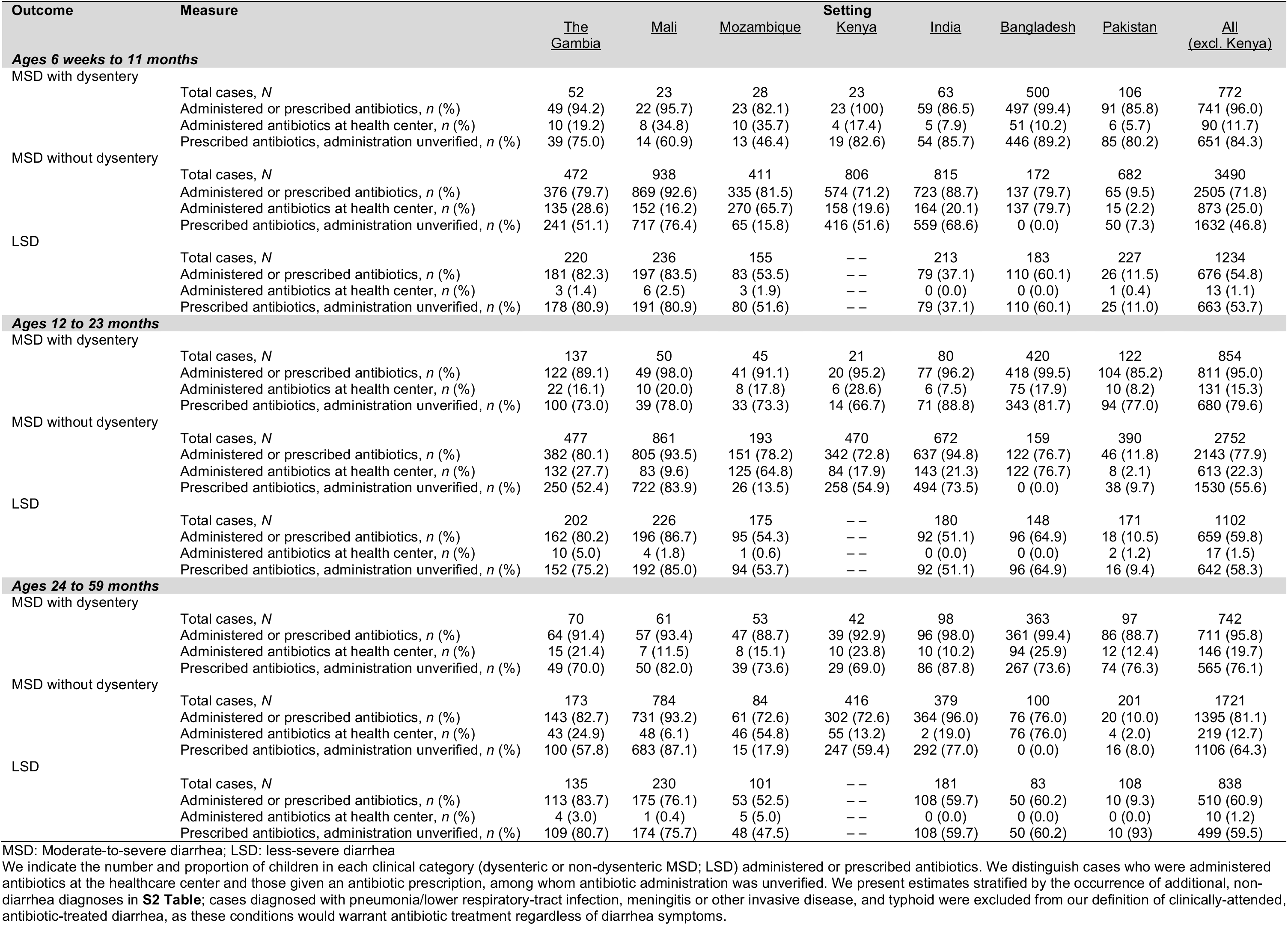
Antibiotic treatment of moderate-to-severe diarrhea and less-severe diarrhea.

Predominant antibiotic choices varied by setting (**Figure 1**). Trimethoprim/sulfamethoxazole was the most common treatment in African sites for both MSD and LSD, while quinolones were more commonly used for MSD in the South Asian sites. Nearly all antibiotic-treated LSD cases in Bangladesh received azithromycin, whereas this drug was less commonly used in other settings.

**Figure 1:**
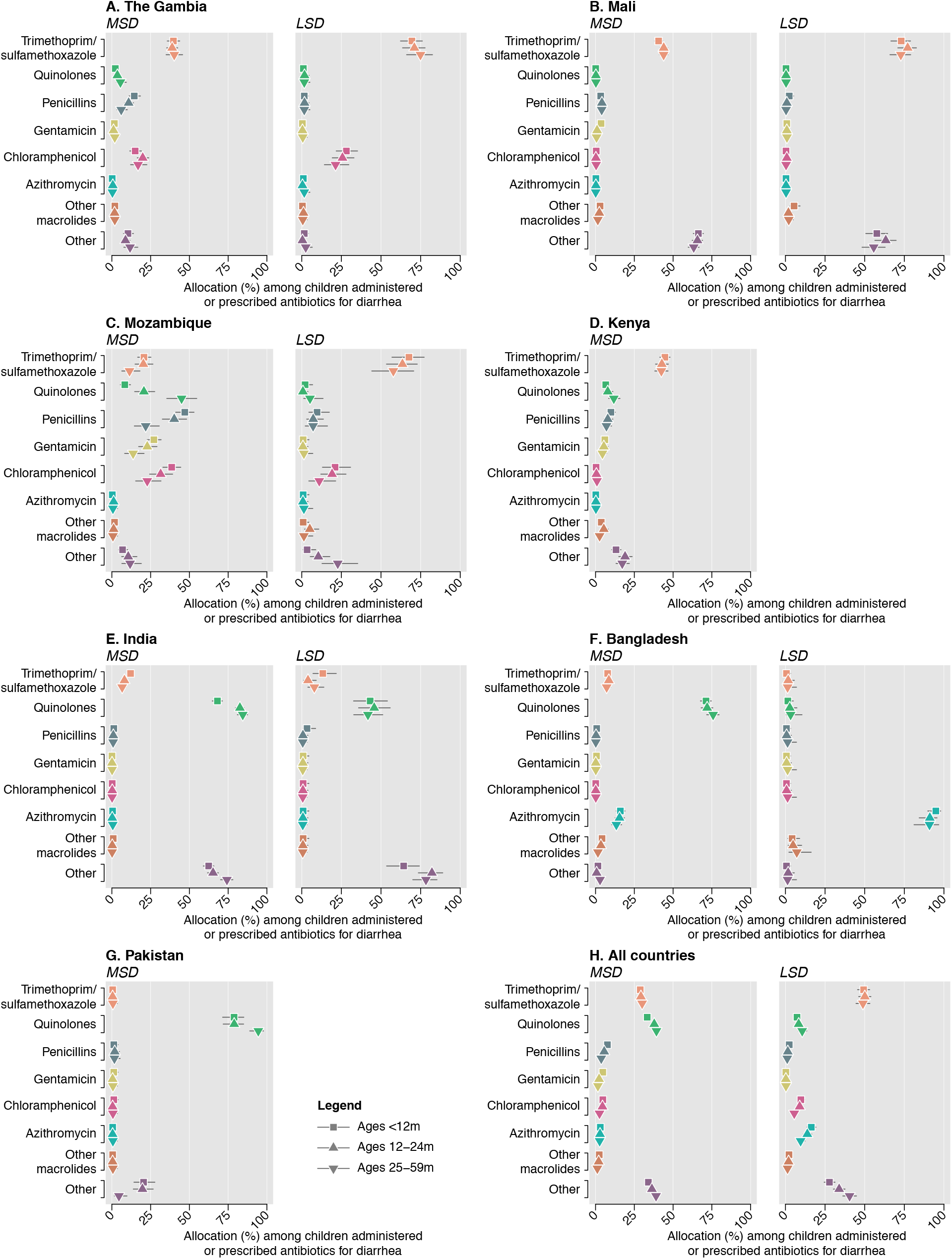
Antibiotics administered and prescribed. We illustrate the proportion of clinically-attended, antibiotic-treated diarrhea cases (stratified as moderate-to-severe diarrhea [MSD] and less-severe diarrhea [LSD] cases) receiving various classes of antibiotics. Proportions are calculated among all cases administered or prescribed an antibiotic who were not diagnosed with pneumonia/lower respiratory tract infection, meningitis or other invasive infection, or typhoid. Totals may not sum to 100%, as each case could receive multiple antibiotics. Estimates are not presented for LSD in Pakistan due to low counts (47 antibiotic-treated cases, total). Data on LSD cases were not collected in Kenya. Lines denote 95% confidence intervals around point estimates.

### Association of pathogen detection with antibiotic treatment

Across all sites, adjusted odds of antibiotic administration or prescribing were higher in *Shigella-positive* than *Shigella-negative* MSD cases not diagnosed with other conditions that would warrant antibiotic treatment (**Figure 2**). In agreement with this finding, *Shigella* detection was associated with higher odds of dysentery among MSD cases. Within the South Asian GEMS study sites, *Shigella* was the only pathogen associated with higher odds of antibiotic administration or prescription, whereas detections of rotavirus, ST-ETEC, or *V. cholerae* O1 among MSD cases each predicted lower odds of antibiotic administration or prescription. Detections of these pathogens among MSD cases were also associated with lower odds of dysentery and higher odds of hospitalization. However, within the African sites, rotavirus-positive MSD cases were more likely than rotavirus-negative cases to be administered or prescribed antibiotics.

**Figure 2:**
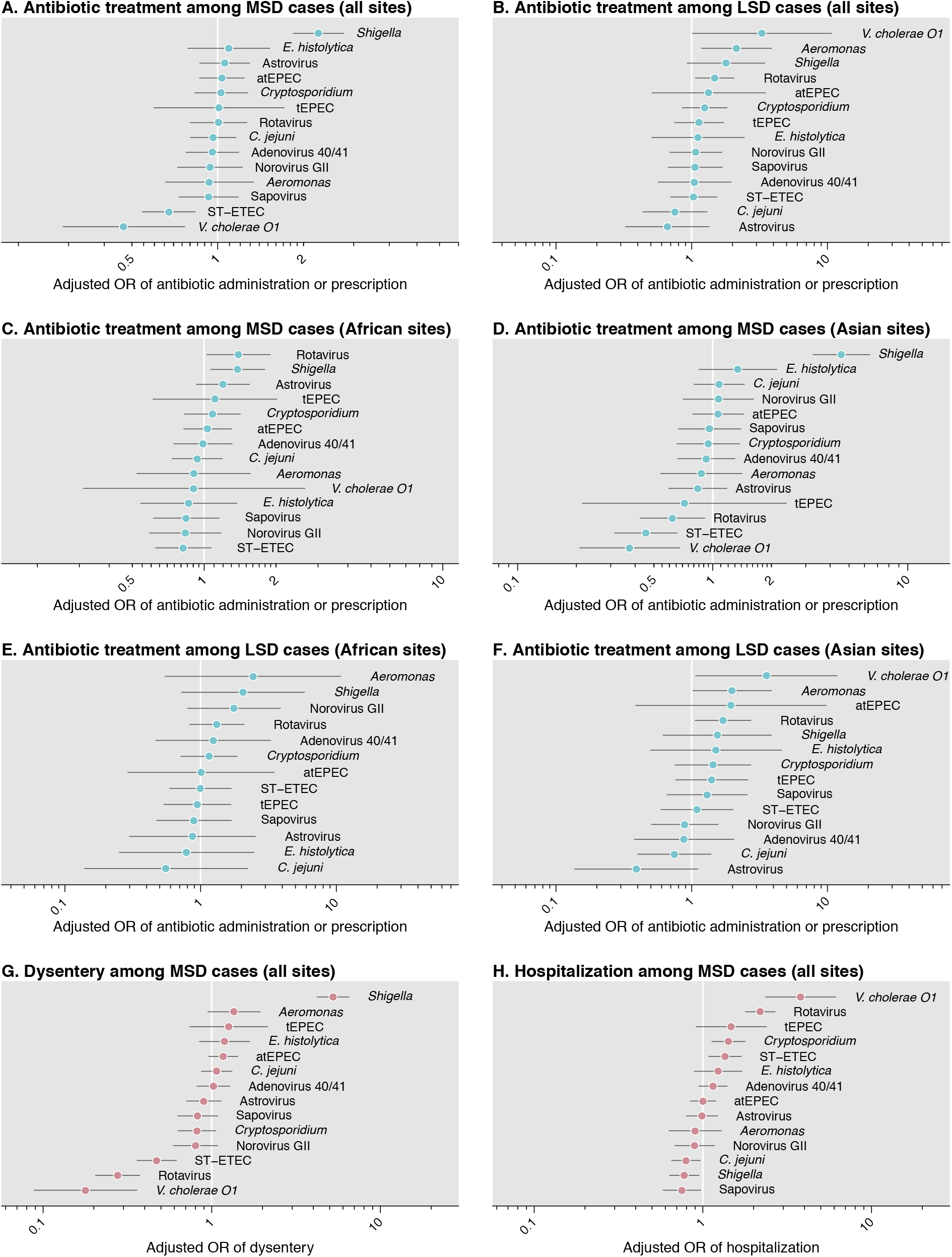
Adjusted association of pathogen with antibiotic prescribing, dysentery, and hospitalization. We illustrate adjusted odds ratios for the association of each pathogen with antibiotic treatment among (**A**) MSD cases, and (**B**), LSD cases. We next stratify estimates for the association of pathogen with antibiotic receipt, among MSD cases, by (**C**) sub-Saharan African sites and (**D**) South Asian sites, and among LSD cases, by (**E**) sub-Saharan African sites and (**D**) South Asian sites. Last, we illustrate the association of each pathogen with (**G**) dysentery and (**H**) hospitalization, among MSD cases. Analyses of pathogen presence in MSD define presence as Cq<35 and absence as Cq≥35. Adjusted odds ratios are computed with conditional logistic regression models defining strata by country, age (in months), and period (in fortnights); all models adjust for presence of other pathogens and occurrence of any non-diarrhea diagnosis. Children diagnosed with other conditions justifying antibiotic treatment (lower respiratory tract infection, meningitis or other invasive infection, and typhoid) are excluded from analyses. Lines indicate 95% confidence intervals around (median) point estimates.

Among LSD cases, detections of *V. cholerae* O1, *Aeromonas, Shigella*, and rotavirus were each associated with elevated odds of antibiotic administration or prescribing (**Figure 2**). These findings persisted in analyses stratified by region. In contrast to our observations in MSD, detections of rotavirus were associated with higher odds of antibiotic administration or prescribing in LSD in South Asian sites as well as African sites.

### Etiology of antibiotic-treated diarrhea

Rotavirus was the leading clinically-attended, antibiotic-treated diarrhea etiology overall and during both the first and second years of life, accounting for 34.2% (95% confidence interval: 28.0-43.3%) and 28.8% (22.9-37.2%) of cases in these age groups, respectively (**Tables 2-3**). Rotavirus was associated with the greatest share of LSD cases resulting in antibiotic treatment across all ages, and with the greatest share of MSD resulting in antibiotic treatment during the first year of life (**S5 Table**; **S6 Table**). Although a minor contributor to clinically-attended, antibiotic-treated diarrhea during the first year of life, *Shigella* accounted for 23.4% (17.9-29.7%) and 23.7% (15.3-34.2%) of cases at ages 12-23 months and 24-59 months, respectively, and was thus the second-leading and leading etiology of antibiotic-treated cases at these ages (**Table 2**). In contrast to rotavirus, *Shigella* was associated with >4-fold greater fractions of clinically-attended, antibiotic-treated MSD cases than LSD cases, and was the leading cause of clinically-attended, antibiotic-treated MSD at ages 12-23 months as well as 24-59 months (**S5 Table**; **S6 Table**). In total, we estimated that 14.9% (11.4-18.9%) of clinically-attended, antibiotic-treated diarrhea was attributable to *Shigella* (**Table 3**).

**Table 2:** Adjusted attributable fractions of all clinically-attended, antibiotic-treated diarrhea associated with individual pathogens, by age stratum and site.

| Pathogen | The Gambia | Mali | Mozambique | Setting<br>India | Bangladesh | Pakistan | All (excl. Kenya)<sup>1</sup> |
| --- | --- | --- | --- | --- | --- | --- | --- |
| **Ages 6 weeks to 11 months** |  |  |  |  |  |  |  |
| Rotavirus | 26.5 (19.0, 35.1) | 31.4 (24.4, 37.5) | 50.8 (37.1, 63.5) | 29.6 (21.2, 37.2) | 47.3 (35.3, 64.9) | 14.3 (3.0, 30.2) | 34.2 (28.0, 43.3) |
| Adenovirus 40/41 | 13.0 (2.4, 29.0) | 20.3 (11.1, 29.8) | 36.4 (16.0, 80.0) | 24.2 (15.5, 33.4) | 38.9 (16.4, 80.4) | 42.0 (1.3, 85.5) | 23.1 (0.0, 43.3) |
| Sapovirus | 28.9 (0.4, 88.8) | 15.1 (2.1, 61.1) | 10.4 (0.2, 74.3) | 17.7 (2.0, 51.8) | 5.6 (0.5, 54.5) | 53.6 (1.4, 86.2) | 21.4 (1.2, 63.1) |
| ST-EPEC (ST-only or LT/ST) | 11.9 (6.0, 22.6) | 7.9 (3.9, 13.1) | 16.0 (8.0, 30.5) | 4.8 (2.1, 8.6) | 4.7 (1.7, 12.2) | 22.5 (6.9, 49.7) | 10.3 (5.9, 17.5) |
| Norovirus GII | 14.5 (6.3, 41.8) | 6.5 (3.4, 13.0) | 4.4 (0.8, 12.4) | 5.7 (2.6, 12.9) | 13.2 (3.1, 45.6) | 5.3 (0.3, 25.9) | 9.1 (4.2, 24.6) |
| *Cryptosporidium* | 11.2 (7.8, 16.7) | 14.8 (8.9, 21.6) | 15.6 (9.7, 22.1) | 4.8 (2.7, 7.8) | 2.5 (0.8, 5.8) | 8.5 (0.0, 22.9) | 8.7 (0.0, 13.3) |
| *C. jejuni* | 1.4 (0.5, 3.4) | 4.6 (1.5, 9.7) | 4.2 (0.9, 12.8) | 6.9 (3.5, 11.3) | 4.3 (1.0, 10.4) | 3.8 (1.2, 9.1) | 4.1 (1.9, 6.7) |
| *Shigella* | 2.6 (1.0, 5.8) | 3.0 (1.1, 6.5) | 0.9 (0.0, 4.5) | 3.2 (0.5, 6.6) | 8.8 (2.2, 17.5) | 13.2 (5.6, 29.0) | 3.7 (2.0, 6.1) |
| Astrovirus | 0.3 (0.0, 1.2) | 2.1 (0.6, 5.1) | 0.2 (0.0, 1.2) | 2.8 (1.1, 5.5) | 1.1 (0.2, 3.4) | 0.7 (0.0, 3.0) | 1.3 (0.5, 2.7) |
| *Aeromonas* | 0.0 (0.0, 0.2) | 0.1 (0.0, 0.6) | 0.8 (0.1, 2.4) | 1.2 (0.3, 2.8) | 1.5 (0.3, 3.9) | 0.8 (0.0, 3.7) | 0.7 (0.3, 1.7) |
| tEPEC | 0.2 (0.0, 1.0) | 1.3 (0.1, 4.1) | 0.5 (0.0, 2.9) | 0.7 (0.0, 2.3) | 0.1 (0.0, 1.1) | 0.2 (0.0, 1.7) | 0.6 (0.0, 1.8) |
| *Vibrio cholerae* O1 | — | 0.2 (0.0, 0.7) | — | 0.9 (0.1, 2.1) | 1.5 (0.2, 5.9) | 1.7 (0.0, 11.9) | 0.6 (0.2, 1.9) |
| Non-typhoidal *Salmonella* | 0.1 (0.0, 0.6) | 0.6 (0.0, 1.8) | 0.6 (0.0, 2.3) | 0.0 (0.0, 0.2) | 0.2 (0.0, 1.1) | 0.0 (0.0, 0.3) | 0.2 (0.1, 0.6) |
| *Entamoeba histolytica* | — | 0.0 (0.0, 0.7) | 0.1 (0.0, 4.7) | 0.1 (0.0, 1.6) | 0.4 (0.0, 2.5) | — | 0.1 (0.0, 1.5) |
| **Ages 12 to 23 months** |  |  |  |  |  |  |  |
| Rotavirus | 16.9 (11.3, 24.6) | 19.2 (14.2, 24.5) | 31.5 (21.6, 44.7) | 29.9 (21.9, 36.9) | 63.6 (48.1, 83.8) | 17.1 (1.0, 43.0) | 28.8 (22.9, 37.2) |
| *Shigella* | 20.2 (12.0, 29.5) | 21.4 (7.8, 34.9) | 21.3 (10.4, 35.1) | 18.5 (11.8, 24.6) | 38.9 (20.2, 59.7) | 55.4 (21.9, 92.7) | 23.4 (17.9, 29.7) |
| Sapovirus | 12.2 (0.3, 76.2) | 7.6 (0.7, 50.3) | 13.2 (0.2, 81.8) | 9.3 (1.1, 50.9) | 15.1 (0.6, 70.7) | 10.4 (0.4, 79.7) | 13.3 (0.8, 60.9) |
| Adenovirus 40/41 | 9.8 (2.4, 24.2) | 12.0 (4.2, 20.4) | 23.3 (2.7, 67.9) | 15.1 (8.0, 24.3) | 21.7 (7.8, 51.4) | 2.6 (0.8, 6.4) | 12.4 (0.0, 29.4) |
| ST-EPEC (ST-only or LT/ST) | 13.5 (8.1, 20.8) | 10.1 (6.5, 15.1) | 18.9 (11.6, 29.4) | 9.9 (6.3, 15.3) | 4.8 (2.1, 9.8) | 7.2 (0.8, 20.9) | 11.9 (8.1, 17.3) |
| *Cryptosporidium* | 8.8 (4.9, 16.8) | 6.8 (3.7, 11.0) | 8.6 (1.1, 18.1) | 4.8 (2.7, 7.6) | 1.3 (0.3, 3.1) | 7.6 (1.0, 22.6) | 5.5 (0.0, 10.8) |
| Norovirus GII | 4.1 (1.2, 17.6) | 1.1 (0.2, 4.9) | 2.8 (0.3, 12.6) | 5.8 (2.1, 21.0) | 4.4 (1.3, 21.6) | 7.9 (0.6, 45.8) | 4.4 (1.5, 17.8) |
| *C. jejuni* | 1.0 (0.1, 3.2) | 3.1 (0.1, 10.1) | 1.1 (0.1, 4.2) | 5.0 (0.3, 11.2) | 1.0 (0.0, 3.3) | 2.0 (0.1, 6.4) | 2.4 (0.1, 5.5) |
| Astrovirus | 0.6 (0.1, 1.7) | 2.3 (0.4, 5.8) | 0.3 (0.0, 1.4) | 2.3 (0.8, 4.6) | 1.1 (0.2, 3.1) | 1.7 (0.3, 5.3) | 1.4 (0.5, 2.5) |
| *Vibrio cholerae* O1 | — | 0.1 (0.0, 0.7) | 0.2 (0.0, 1.3) | 3.0 (1.2, 5.4) | 0.2 (0.0, 0.8) | 4.4 (0.0, 15.7) | 1.2 (0.4, 2.4) |
| Non-typhoidal *Salmonella* | 0.7 (0.2, 1.9) | 1.6 (0.4, 3.9) | 0.7 (0.0, 2.8) | 0.3 (0.0, 0.9) | 0.6 (0.1, 1.8) | — | 0.7 (0.2, 1.3) |
| *Entamoeba histolytica* | — | 0.0 (0.0, 0.9) | 0.7 (0.0, 10.9) | 0.5 (0.0, 3.5) | 1.7 (0.1, 12.3) | 0.3 (0.0, 1.8) | 0.5 (0.0, 4.3) |
| *Aeromonas* | 0.0 (0.0, 0.2) | 0.4 (0.0, 1.5) | 0.5 (0.0, 2.4) | 0.5 (0.0, 2.1) | 0.7 (0.0, 3.2) | 1.1 (0.0, 5.3) | 0.5 (0.0, 1.5) |
| tEPEC | 0.1 (0.0, 0.8) | 0.1 (0.0, 1.2) | 0.0 (0.0, 0.5) | 0.0 (0.0, 0.6) | 0.0 (0.0, 0.2) | 0.0 (0.0, 0.4) | 0.0 (0.0, 0.5) |
| **Ages 24 to 59 months** |  |  |  |  |  |  |  |
| *Shigella* | 24.7 (14.5, 35.1) | 11.0 (1.7, 23.6) | 28.2 (10.3, 48.4) | 14.9 (4.6, 27.1) | 78.4 (46.6, 87.7) | 19.6 (6.6, 40.8) | 23.7 (15.3, 34.2) |
| Rotavirus | 12.0 (6.0, 19.5) | 9.4 (4.0, 15.0) | 34.0 (16.0, 61.2) | 17.2 (9.9, 25.4) | 18.6 (9.6, 48.1) | 0.8 (0.1, 2.3) | 17.7 (10.9, 29.9) |
| ST-EPEC (ST-only or LT/ST) | 9.8 (4.8, 19.0) | 2.8 (1.1, 5.6) | 11.0 (4.8, 21.9) | 3.0 (1.0, 5.8) | 2.5 (0.5, 10.1) | 13.4 (3.0, 34.8) | 6.3 (3.2, 12.4) |
| Norovirus GII | 3.4 (0.9, 9.1) | 3.1 (1.1, 7.2) | 1.2 (0.0, 5.9) | 4.1 (1.6, 9.6) | 5.6 (1.8, 22.0) | 1.2 (0.2, 3.8) | 3.6 (1.5, 7.6) |
| *Vibrio cholerae* O1 | 0.3 (0.0, 1.2) | 0.3 (0.0, 1.2) | 2.4 (0.3, 9.9) | 6.2 (2.1, 10.9) | 1.8 (0.3, 4.0) | 12.6 (1.5, 38.1) | 3.5 (1.6, 7.2) |
| *C. jejuni* | 1.4 (0.1, 4.4) | 4.6 (0.5, 13.9) | 0.7 (0.0, 3.8) | 3.9 (0.6, 9.9) | 0.9 (0.1, 2.5) | 3.7 (0.5, 10.8) | 2.8 (0.6, 6.6) |
| Adenovirus 40/41 | 3.4 (0.1, 18.4) | 2.1 (0.1, 9.5) | 6.1 (0.1, 38.2) | 4.9 (0.3, 17.1) | 9.4 (0.3, 28.3) | 0.9 (0.0, 3.5) | 2.6 (0.0, 15.4) |
| Sapovirus | 1.3 (0.4, 5.5) | 1.9 (0.3, 8.5) | 1.6 (0.2, 6.7) | 3.0 (0.7, 13.0) | 4.4 (0.8, 13.2) | 1.8 (0.4, 4.8) | 2.3 (1.1, 8.6) |
| *Entamoeba histolytica* | — | 2.0 (0.4, 5.8) | 0.3 (0.0, 7.1) | 1.4 (0.3, 5.8) | 0.5 (0.0, 5.8) | 2.9 (0.7, 7.7) | 1.2 (0.4, 4.9) |
| Astrovirus | 0.9 (0.1, 2.9) | 1.0 (0.1, 3.3) | 0.8 (0.0, 4.6) | 1.0 (0.2, 2.5) | 2.2 (0.3, 5.3) | 2.1 (0.3, 6.1) | 1.2 (0.3, 2.4) |
| *Aeromonas* | — | 0.1 (0.0, 0.9) | 1.2 (0.1, 4.2) | 1.2 (0.2, 3.3) | 1.5 (0.2, 3.9) | 3.0 (0.1, 11.7) | 1.0 (0.2, 2.6) |
| Non-typhoidal *Salmonella* | 1.1 (0.3, 3.1) | 1.3 (0.2, 3.9) | 0.4 (0.0, 2.5) | 0.4 (0.1, 1.3) | 0.3 (0.0, 1.4) | 1.2 (0.0, 4.2) | 0.8 (0.3, 1.5) |
| *Cryptosporidium* | 2.4 (0.0, 69.1) | 1.5 (0.0, 36.2) | 2.2 (0.0, 58.0) | 2.1 (0.3, 47.5) | 0.2 (0.0, 27.5) | 0.3 (0.0, 1.5) | 0.5 (0.0, 37.8) |
| tEPEC | 0.3 (0.0, 1.4) | 0.3 (0.0, 2.2) | 0.2 (0.0, 2.1) | 0.2 (0.0, 1.3) | 0.1 (0.0, 0.8) | 0.2 (0.0, 2.0) | 0.3 (0.0, 1.2) |
| atEPEC | — | 0.0 (0.0, 0.1) | — | 0.1 (0.0, 0.6) | — | — | 0.0 (0.0, 0.2) |

Abbreviations: ST-EPEC (ST-only or LT/ST): Toxigenic *E. coli* encoding Shiga toxin (stable toxin or heat-labile toxin); tEPEC: Typical enteropathogenic *E. coli*; atEPEC: Atypical enteropathogenic *E. coli*. Estimates are presented as adjusted attributable fractions (%) of age-specific clinically-attended, antibiotic-treated diarrhea cases for each pathogen, together with 95% confidence intervals, controlling for presence of the other pathogens and their abundance (in MSD cases). We obtain estimates by summing pathogen-attributable incidence of cases with moderate-to-severe and less-severe symptoms (as estimated in **S5 Table S5** and **S6 Table**). Empty cells indicate where point estimates of the model-estimated adjusted attributable fraction were zero or below, indicating no statistical association with the clinically-attended, antibiotic-treated diarrhea endpoint. We present estimates based on conventional diagnostic data in **S9 Table**. Estimated incidence rates exclude diarrhea cases receiving antibiotics who were also diagnosed with pneumonia/lower respiratory tract infection, meningitis or other invasive disease, or typhoid, as these conditions warrant antibiotic treatment irrespective of diarrhea symptoms.
1. Aggregated estimates across sites weight individual cases so that each site receives equal weight within each age stratum.

**Table 3:** Adjusted attributable fractions of all clinically-attended, antibiotic-treated diarrhea associated with individual pathogens, by site.

| Pathogen | Setting |  |  |  |  |  |  |
| --- | --- | --- | --- | --- | --- | --- | --- |
|  | The Gambia | Mali | Mozambique | India | Bangladesh | Pakistan | All (excl. Kenya)<sup>1</sup> |
| Rotavirus | 20.5 (15.7, 26.7) | 22.2 (16.6, 28.0) | 40.6 (30.9, 52.5) | 26.5 (21.5, 31.5) | 49.8 (36.6, 64.0) | 12.2 (3.8, 24.7) | 29.2 (24.5, 35.2) |
| Adenovirus 40/41 | 10.5 (4.3, 20.4) | 13.2 (7.4, 21.2) | 29.6 (11.3, 60.7) | 16.2 (10.8, 22.8) | 28.7 (13.5, 62.8) | 24.5 (1.7, 48.6) | 16.3 (0.0, 29.6) |
| *Shigella* | 12.5 (7.9, 18.4) | 10.3 (5.1, 16.9) | 13.3 (6.0, 23.2) | 11.6 (7.5, 16.7) | 30.5 (13.5, 49.0) | 28.6 (15.5, 48.6) | 14.9 (11.4, 18.9) |
| Sapovirus | 19.5 (0.6, 69.4) | 10.7 (1.8, 45.3) | 10.9 (0.5, 60.5) | 11.8 (2.2, 35.8) | 9.8 (1.1, 49.1) | 27.9 (1.5, 64.1) | 14.6 (1.4, 48.2) |
| ST-EPEC (ST-only or LT/ST) | 12.4 (8.1, 18.9) | 7.4 (4.8, 11.2) | 16.2 (10.7, 25.7) | 6.3 (4.2, 9.2) | 4.8 (2.5, 9.7) | 15.7 (6.7, 31.1) | 10.1 (7.4, 14.4) |
| *Cryptosporidium* | 9.8 (6.6, 19.5) | 9.5 (5.9, 17.6) | 11.5 (5.3, 21.1) | 4.5 (2.8, 13.9) | 2.0 (0.7, 9.0) | 7.6 (1.6, 15.3) | 6.5 (0.0, 13.6) |
| Norovirus GII | 9.1 (4.3, 28.4) | 4.1 (2.2, 8.7) | 3.3 (1.2, 10.4) | 5.5 (3.3, 12.2) | 9.6 (3.6, 31.9) | 5.7 (1.2, 23.2) | 6.4 (3.6, 16.8) |
| *C. jejuni* | 1.3 (0.5, 2.4) | 4.1 (1.7, 8.0) | 2.1 (0.8, 4.9) | 5.6 (3.0, 8.7) | 2.4 (0.9, 5.1) | 3.2 (1.5, 6.1) | 3.2 (1.9, 5.1) |
| *V. cholerae* O1 | 0.0 (0.0, 0.2) | 0.2 (0.0, 0.6) | 0.5 (0.1, 1.9) | 3.0 (1.7, 5.2) | 1.1 (0.3, 4.0) | 5.4 (1.2, 15.0) | 1.5 (0.8, 2.6) |
| Astrovirus | 0.5 (0.2, 1.1) | 1.9 (0.8, 3.8) | 0.4 (0.1, 1.3) | 2.2 (1.1, 3.7) | 1.3 (0.4, 2.7) | 1.4 (0.5, 3.2) | 1.3 (0.7, 2.1) |
| *Aeromonas* | 0.0 (0.0, 0.1) | 0.2 (0.0, 0.7) | 0.8 (0.2, 1.8) | 1.0 (0.4, 2.0) | 1.3 (0.4, 3.1) | 1.6 (0.3, 4.7) | 0.7 (0.3, 1.4) |
| *E. histolytica* | — | 0.5 (0.1, 2.0) | 0.6 (0.0, 8.7) | 0.6 (0.2, 3.9) | 1.0 (0.2, 6.4) | 0.8 (0.2, 1.9) | 0.5 (0.2, 3.5) |
| Non-typhoidal *Salmonella* | 0.5 (0.2, 1.1) | 1.1 (0.4, 2.2) | 0.6 (0.2, 1.5) | 0.2 (0.0, 0.6) | 0.4 (0.1, 1.0) | 0.3 (0.0, 0.9) | 0.5 (0.3, 0.8) |
| tEPEC | 0.2 (0.0, 0.6) | 0.7 (0.1, 2.1) | 0.3 (0.0, 1.1) | 0.4 (0.0, 1.1) | 0.1 (0.0, 0.5) | 0.2 (0.0, 0.9) | 0.4 (0.0, 0.9) |

Abbreviations: ST-EPEC (ST-only or LT/ST): Toxigenic *E. coli* encoding Shiga toxin (stable toxin or heat-labile toxin); tEPEC: Typical enteropathogenic *E. coli*; atEPEC: Atypical enteropathogenic *E. coli*.
1. Aggregated estimates across sites weight individual cases so that each site receives equal weight within each age stratum. Estimated incidence rates exclude diarrhea cases receiving antibiotics who were also diagnosed with pneumonia/lower respiratory tract infection, meningitis or other invasive disease, or typhoid, as these conditions warrant antibiotic treatment irrespective of diarrhea symptoms.

Adenovirus serotypes 40/41, sapovirus, and ST-ETEC were also important etiologies of antibiotic-treated diarrhea during the first year of life, accounting for 23.1% (0.0-43.3%), 21.4% (1.2-63.1%), and 10.3% (5.9-17.5%) of cases in this age group, respectively (**Table 2**). Of these pathogens, only ST-ETEC was a prominent cause of clinically-attended, antibiotic-treated diarrhea across all ages; we estimated that ST-ETEC accounted for 12.4% (0.0-29.4%) and 6.3% (3.2-12.4%) of cases at ages 12-23 months and 24-59 months, respectively. In contrast, by ages 24-59 months, adenovirus 40/41 and sapovirus each accounted for <3% of all clinically-attended, antibiotic-treated diarrhea.

We noted several regional differences in etiologies of clinically-attended, antibiotic-treated diarrhea. Overall and within most age groups, *Shigella* was associated with roughly 2-fold greater fractions of cases in Bangladesh and Pakistan as compared to other sites (**Tables 2-3**). Additionally, *V. cholerae* O1 accounted for 3.3% (1.7-5.2%) and 5.4% (1.2-15.0%) of clinically-attended, antibiotic-treated diarrhea in India and Pakistan, respectively, but <1% of cases in each African site (**Table 3**). In contrast, estimates of the etiologic fraction for *Cryptosporidium* tended to be larger in the African sites than the South Asian sites.

### Appropriate and inappropriate antibiotic treatment

We estimated that dysentery occurred in only 17.5% (11.2-22.5%) of clinically-attended, antibiotic-treated diarrhea cases across all settings (**Table 4**). Whereas 52.4% (19.5-70.4%) and 24.5% (13.2-41.5%) of incident clinically-attended, antibiotic-treated diarrhea cases in Bangladesh and Pakistan experienced dysentery, the fraction was below 10% in all other settings. On this basis, the ratio of inappropriate to appropriate treatment of diarrhea cases with antibiotics ranged from 0.9 (0.4-4.1) in Bangladesh to 29.8 (16.6-75.3) in Mali.

**Table 4:**
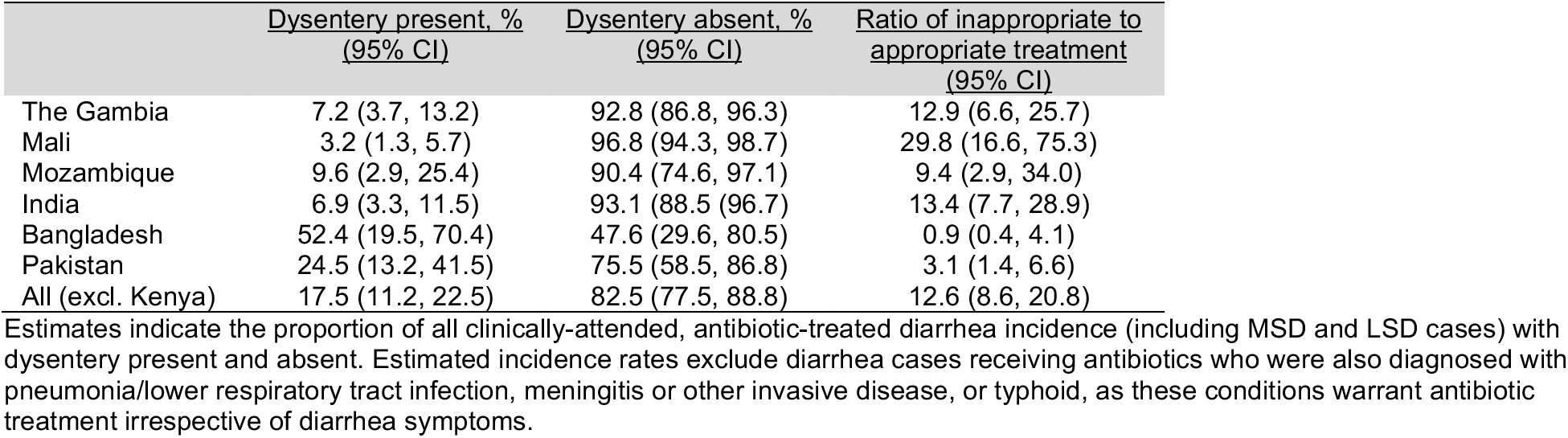
Appropriate and inappropriate antibiotic treatment based on occurrence of dysentery, by site.

|  | <u>Dysentery present. %</u><br><u>(95% CI)</u> | <u>Dysentery absent. %</u><br><u>(95% CI)</u> | <u>Ratio of inappropriate to</u><br><u>appropriate treatment</u><br><u>(95% CI)</u> |
| --- | --- | --- | --- |
| The Gambia | 7.2 (3.7, 13.2) | 92.8 (86.8, 96.3) | 12.9 (6.6, 25.7) |
| Mali | 3.2 (1.3, 5.7) | 96.8 (94.3, 98.7) | 29.8 (16.6, 75.3) |
| Mozambique | 9.6 (2.9, 25.4) | 90.4 (74.6, 97.1) | 9.4 (2.9, 34.0) |
| India | 6.9 (3.3, 11.5) | 93.1 (88.5, 96.7) | 13.4 (7.7, 28.9) |
| Bangladesh | 52.4 (19.5, 70.4) | 47.6 (29.6, 80.5) | 0.9 (0.4, 4.1) |
| Pakistan | 24.5 (13.2, 41.5) | 75.5 (58.5, 86.8) | 3.1 (1.4, 6.6) |
| All (excl. Kenya) | 17.5 (11.2, 22.5) | 82.5 (77.5, 88.8) | 12.6 (8.6, 20.8) |

Estimates indicate the proportion of all clinically-attended, antibiotic-treated diarrhea incidence (including MSD and LSD cases) with dysentery present and absent. Estimated incidence rates exclude diarrhea cases receiving antibiotics who were also diagnosed with pneumonia/lower respiratory tract infection, meningitis or other invasive disease, or typhoid, as these conditions warrant antibiotic treatment irrespective of diarrhea symptoms.

Non-dysenteric cases accounted for over half of *Shigella-*attributable antibiotic consumption, as well as 15.3% (11.4-20.4%) and 12.2% (7.3-20.1%) of all clinically-attended, antibiotic-treated diarrhea cases at ages 12-23 months and 24-59 months, respectively (**Table 5**). This finding was consistent in all settings except Bangladesh and Pakistan, where the majority of antibiotic-treated, *Shigella*-attributable cases were dysenteric.

**Table 5:** Fraction of clinically-attended, antibiotic-treated diarrhea contributed by dysenteric and non-dysenteric shigellosis.

| Outcome | The Gambia | Mali | Mozambique | Setting<br>India | Bangladesh | Pakistan | All (excl. Kenya)<sup>1</sup> |
| --- | --- | --- | --- | --- | --- | --- | --- |
| **Age 6 weeks to 11 months** |  |  |  |  |  |  |  |
| Dysenteric (MSD only) | 1.3 (0.5, 3.4) | 0.3 (0.0, 1.6) | 0.4 (0.0, 1.7) | 1.2 (0.3, 2.4) | 8.4 (2.1, 16.6) | 12.0 (5.5, 22.4) | 2.3 (1.3, 3.6) |
| Non-dysenteric (MSD and LSD) | 1.2 (0.5, 3.0) | 2.6 (1.0, 5.7) | 0.4 (0.0, 3.7) | 2.1 (0.0, 5.1) | 0.4 (0.0, 1.2) | 0.0 (0.0, 14.2) | 1.3 (0.4, 3.3) |
| All *Shigella*-attributable cases | 2.6 (1.0, 5.8) | 3.0 (1.1, 6.5) | 0.9 (0.0, 4.5) | 3.2 (0.5, 6.6) | 8.8 (2.2, 17.5) | 13.2 (5.6, 29.0) | 3.7 (2.0, 6.1) |
| **Age 12 to 23 months** |  |  |  |  |  |  |  |
| Dysenteric (MSD only) | 5.9 (2.7, 11.4) | 2.2 (0.6, 4.3) | 6.5 (2.3, 16.1) | 4.0 (2.2, 6.5) | 28.7 (10.1, 52.3) | 19.1 (7.8, 37.2) | 7.9 (5.1, 11.9) |
| Non-dysenteric (MSD and LSD) | 14.2 (7.9, 21.1) | 19.1 (7.0, 31.4) | 14.5 (6.4, 23.0) | 14.4 (8.7, 19.5) | 9.1 (4.7, 17.6) | 35.3 (2.8, 77.5) | 15.3 (11.4, 20.4) |
| All *Shigella*-attributable cases | 20.2 (12.0, 29.5) | 21.4 (7.8, 34.9) | 21.3 (10.4, 35.1) | 18.5 (11.8, 24.6) | 38.9 (20.2, 59.7) | 55.4 (21.9, 92.7) | 23.4 (17.9, 29.7) |
| **Age 24 to 59 months** |  |  |  |  |  |  |  |
| Dysenteric (MSD only) | 6.8 (2.6, 14.4) | 2.0 (0.3, 5.0) | 9.4 (1.2, 30.9) | 6.5 (1.6, 12.9) | 58.7 (10.0, 79.8) | 18.8 (6.4, 39.0) | 10.9 (5.6, 21.0) |
| Non-dysenteric (MSD and LSD) | 17.2 (9.0, 27.4) | 8.9 (1.4, 19.3) | 16.9 (5.7, 31.4) | 8.3 (2.6, 15.0) | 17.3 (6.8, 51.8) | 0.8 (0.2, 2.1) | 12.2 (7.3, 20.1) |
| All *Shigella*-attributable cases | 24.7 (14.5, 35.1) | 11.0 (1.7, 23.6) | 28.2 (10.3, 48.4) | 14.9 (4.6, 27.1) | 78.4 (46.6, 87.7) | 19.6 (6.6, 40.8) | 23.7 (15.3, 34.2) |
| **Age 6 weeks to 59 months** |  |  |  |  |  |  |  |
| Dysenteric (MSD only) | 3.8 (2.2, 6.5) | 1.3 (0.5, 2.5) | 4.1 (1.6, 9.4) | 3.5 (2.0, 6.2) | 22.0 (8.8, 41.2) | 15.5 (9.4, 24.2) | 6.1 (4.3, 8.6) |
| Non-dysenteric (MSD and LSD) | 8.5 (5.0, 13.3) | 8.9 (4.3, 14.8) | 8.7 (3.4, 16.4) | 8.0 (5.0, 11.4) | 6.8 (2.4, 19.8) | 12.1 (1.6, 33.2) | 8.6 (6.4, 11.6) |
| All *Shigella*-attributable cases | 12.5 (7.9, 18.4) | 10.3 (5.1, 16.9) | 13.3 (6.0, 23.2) | 11.6 (7.5, 16.7) | 30.5 (13.5, 49.0) | 28.6 (15.5, 48.6) | 14.9 (11.4, 18.9) |

Abbreviations: MSD: moderate-to-severe diarrhea; LSD: less-severe diarrhea.
1. Aggregated estimates across sites weight individual cases so that each site receives equal weight within each age.

### Incidence of clinically-attended, antibiotic-treated diarrhea

Across sites, we estimated incidence rates of 12.2 (9.0-17.8), 10.2 (7.4-13.9), and 1.9 (1.3-3.0) clinically-attended, antibiotic-treated diarrhea cases per 100 child-years at risk at ages 6 weeks to 11 months, 12–23 months, and 24-59 months, respectively (**Figure 3**). Consistent with this pattern, incidence rates declined with older age within each setting. Overall and within each age group, incidence rates were highest in India and lowest in Pakistan, where the fewest cases received antibiotics (**Table 1**).

**Figure 3:**
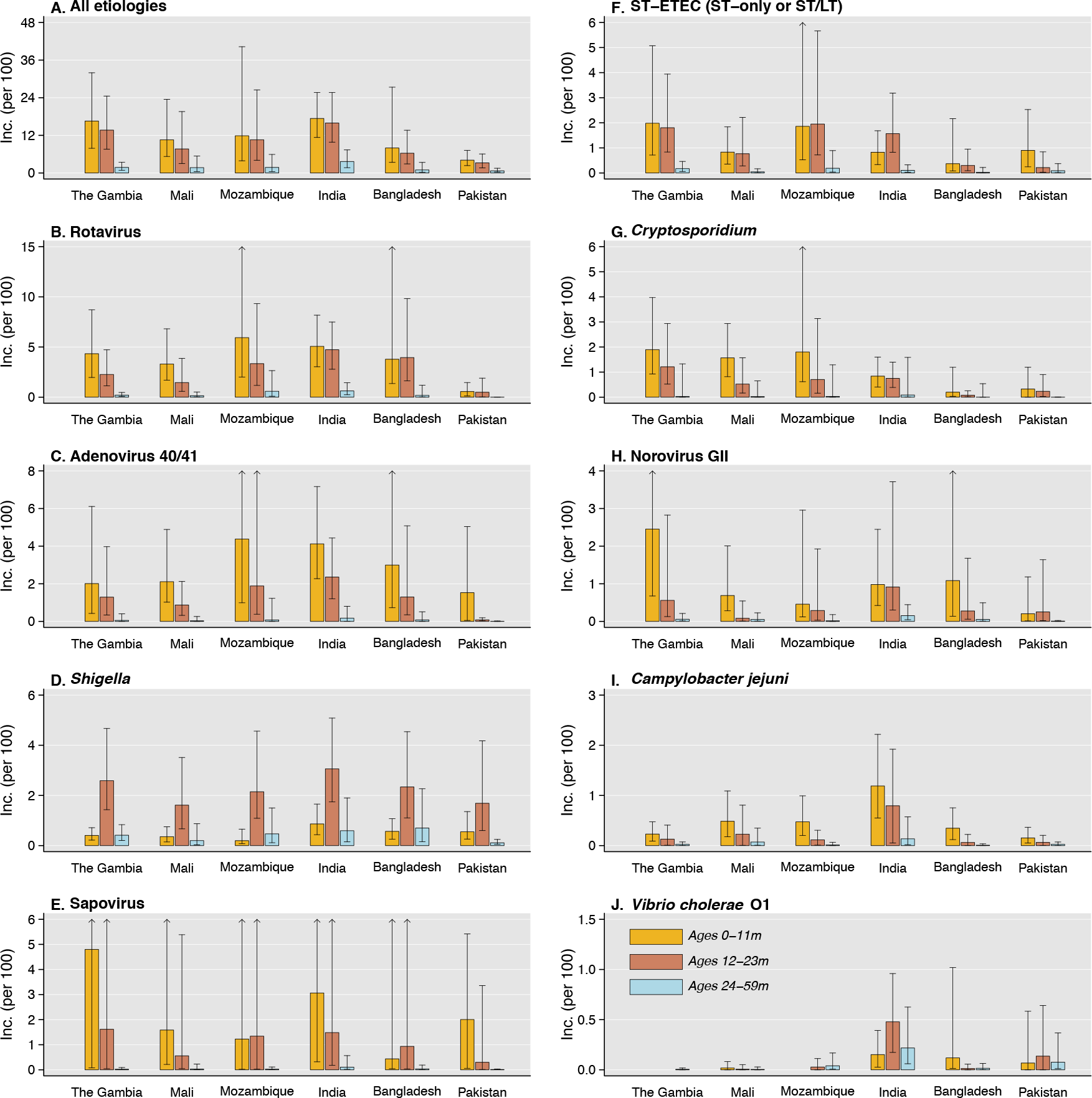
Pathogen-attributable incidence of clinically-attended, antibiotic-treated diarrhea, by site and age stratum. We illustrate age-specific incidence of clinically-attended, antibiotic-treated diarrhea estimates per 100 children annually, according to etiology: (**A**) all causes; (**B**) rotavirus; (**C**) adenovirus serotypes 40/41; (**D**) *Shigellla;* (**E**) sapovirus (**F**) ST-ETC; (**G**) *Cryptosporidium;* (**H**) norovirus GII; (**I**) *C. jejuni;* and (**J**) *V. cholerae* O1. Incidence rates are estimated as the summed rates of incidence of all-cause clinically-attended, antibiotic treated MSD and LSD (**S3 Table**) multiplied by site-specific adjusted attributable fraction estimates for each pathogen in each of these syndromes (**Table 2**; **S5 Table**; **S6 Table**). Lines denote 95% confidence intervals around median rate estimates (bars). Arrows indicate where upper confidence bounds exceeded the plotted range. Estimated incidence rates exclude diarrhea cases receiving antibiotics who were also diagnosed with pneumonia/lower respiratory tract infection, meningitis or other invasive disease, or typhoid, as these conditions warrant antibiotic treatment irrespective of diarrhea symptoms.

Incidence rates of clinically-attended, antibiotic-treated diarrhea attributable to rotavirus, adenovirus 40/41, *Cryptosporidium*, norovirus GII, and *Campylobacter jejuni* declined with older age in each setting (**Figure 3**). A similar pattern appeared in age-specific incidence of clinically-attended, antibiotic-treated diarrhea attributable to sapovirus and ST-ETEC, albeit less consistently. In contrast, *Shigella-attributable* incidence was highest among children ages 12-23 months in all settings. Similarly, incidence attributable to *Vibrio cholerae* O1 peaked at ages 12-23 months in India and Pakistan, where burden of this pathogen was greatest.

Within each age group, LSD cases accounted for the majority of clinically-attended, antibiotic-treated diarrhea incidence (**Figure 4**). However, the relative contributions of MSD and LSD varied by pathogen. Point estimates suggested rotavirus, adenovirus 40/41, sapovirus, ST-ETEC, *Cryptosporidium*, and norovirus GII were each associated with a greater share of less-severe than moderate-to-severe episodes of clinically-attended, antibiotic-treated diarrhea. In contrast, we estimated that *Shigella* caused higher incidence of moderate-to-severe than less-severe cases overall and within each age group.

**Figure 4:**
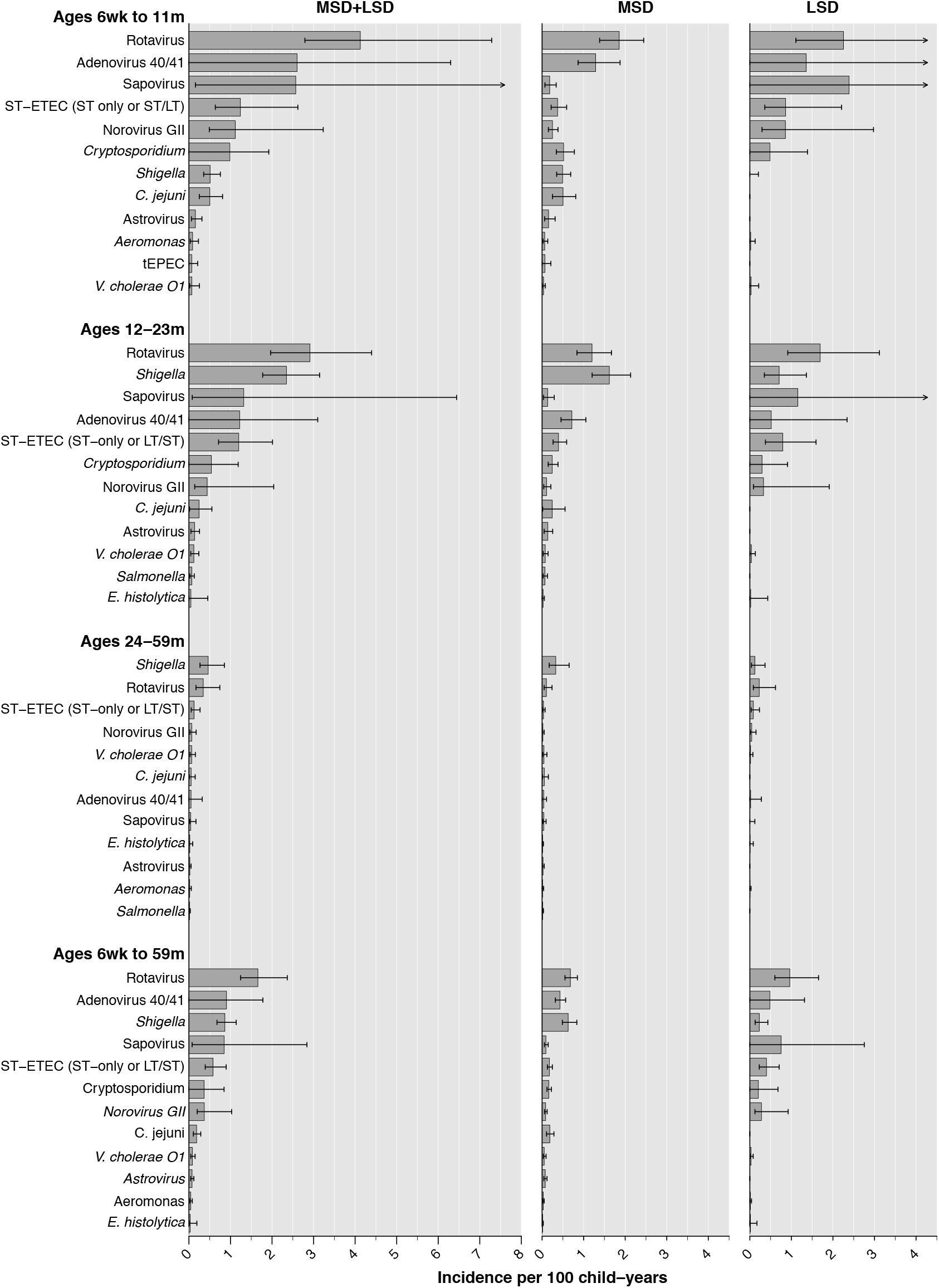
Incidence of clinically-attended, antibiotic-treated diarrhea attributable to enteric pathogens, by age stratum and severity. We illustrate age-specific incidence of diarrhea episodes resulting in antibiotic treatment per 100 children annually, for the twelve leading etiologies in each age group, aggregated across all sites (excluding Kenya). Incidence rates in the left column are estimated as the summed rates of incidence of all-cause clinically-attended, antibiotic-treated MSD and LSD (**S3 Table**) multiplied by site-specific adjusted attributable fraction estimates for each pathogen (**Table 2**; **S5 Table**; **S6 Table**). The two right columns present estimates for MSD and LSD. Lines denote 95% confidence intervals around median rate estimates (bars). Arrows indicate where upper confidence bounds exceeded the plotted range. Estimated incidence rates exclude diarrhea cases receiving antibiotics who were also diagnosed with pneumonia/lower respiratory tract infection, meningitis or other invasive disease, or typhoid, as these conditions warrant antibiotic treatment irrespective of diarrhea symptoms. Abbreviations: ST-ETEC (ST-only or LT/ST): Toxigenic *E. coli* encoding Shiga toxin (stable toxin or heat-labile toxin); tEPEC: Typical enteropathogenic *E. coli*.

## DISCUSSION

Preventing antibiotic use is an important component of the impact and value proposition of numerous vaccines presently in development or in routine use [24]. Our estimates reveal the incidence of clinically-attended, antibiotic-treated diarrhea among young children in LMICs across sub-Saharan Africa and South Asia, and identify most of this treatment as inappropriate under current guidelines [22,23]. Prior to vaccine implementation, rotavirus accounted for 29% of clinically-attended, antibiotic-treated diarrhea among children in these settings. *Shigella*, for which vaccines are presently in clinical trials [25], accounted for an additional 15% of cases, including 25% of cases among children ages 24-59 months. These findings provide a crucial baseline against which the impact of vaccines on diarrhea-related antibiotic use can be assessed.

Choices of antibiotics varied markedly across settings. Whereas the largest share of MSD cases in South Asian settings received quinolones, most MSD cases in sub-Saharan African settings received trimethoprim-sulfamethoxazole. In accordance with WHO guidelines, these antibiotic choices are likely guided by knowledge of local resistance patterns, including near-universal resistance of *Shigella* isolates in Asia to ampicillin and trimethoprim-sulfamethoxazole [26]. However, emerging quinolone and macrolide resistance in *Shigella* and other pathogens may undermine the effectiveness of antibiotic use patterns identified in GEMS data [27,28]. For instance, increasing macrolide resistance has been reported in *Shigella* as well as *E. coli, S. typhi, V. cholerae* O1 and respiratory bacteria in Bangladesh [29], where nearly all antibiotic-treated LSD cases received azithromycin.

Several differences across settings in the clinical spectrum of diarrhea cases receiving antibiotics should be noted. Nearly all children with dysentery were administered or prescribed antibiotics in each setting. However, antibiotics were also administered or prescribed to the majority of non-dysenteric cases. We estimated a ratio of 9-30 cases of inappropriate antibiotic administration or prescribing (i.e., for non-dysenteric cases) per appropriately-treated dysentery case in The Gambia, Mali, Mozambique, and India. A lower but substantial proportion of antibiotic-treated episodes were non-dysenteric in Bangladesh and Pakistan. Whereas this finding reflected lower rates of inappropriate antibiotic treatment in Pakistan, in Bangladesh this finding was instead driven by high prevalence of dysentery, as the proportion of non-dysenteric MSD and LSD cases receiving antibiotics resembled observations in other settings.

*Shigella* detection was strongly associated with antibiotic administration or prescription in all settings. Left untreated, shigellosis is associated with severe adverse outcomes including mortality and growth faltering, independent of dysentery [30-32]. Because the proportion of non-dysenteric *Shigella* infections exceeds the proportion with dysentery, current guidelines for antibiotic treatment of only dysenteric cases may be overly restrictive [20,32]. However, benefits of a broader antibiotic treatment recommendation must be weighed against the potential for expanding inappropriate or unnecessary antibiotic treatment further beyond the levels revealed in our study. Implementation of an effective vaccine against *Shigella* could offer synergistic benefits, for instance by reducing the incidence of shigellosis cases that drive antibiotic use as well as reducing clinical suspicion of shigellosis and resulting empirical antibiotic treatment of diarrhea attributable to other causes. Moreover, expansion of multidrug-resistant *Shigella* lineages, most notably in South Asia, has resulted in a diminishing number of practical treatment options [25]. The emerging prospect of untreatable shigellosis adds heightened urgency to the development of vaccines against *Shigella*.

While general limitations of GEMS have been addressed previously [33], several considerations are salient for this analysis. First, because GEMS enrolled cases seeking care at sentinel hospitals and health centers, our estimates account only for clinically-attended diarrhea. Antibiotics are available without a prescription in many LMICs, and diarrhea cases treated outside healthcare settings may differ in severity and etiology. Therefore, our assessments of pathogen-specific clinically-attended, antibiotic-treated incidence represent the minimum estimate of antibiotic consumption that could be prevented by vaccines or other pathogen-specific interventions [3]. Second, among cases who were prescribed antibiotics, we do not know whether prescriptions were ultimately filled, or whether children received the specific drugs prescribed by clinicians; we also lack data addressing whether children took antibiotics to treat diarrheal illness before the enrollment visit. Accurate reporting of antibiotic use was not considered feasible in these settings, where antibiotics can be obtained in both the formal and informal sectors. Data from the MAL-ED study—a contemporaneous birth-cohort study of diarrhea in LMICs—may provide complementary insights into antibiotic-treated diarrhea episodes for which care was not sought at a clinic [3,34].

At present, no studies have estimated the impact of programmatic rotavirus vaccination on antibiotic use in either LMICs or high-income countries [12]. Our findings suggest that even a moderately effective rotavirus vaccination program could substantially reduce antibiotic consumption. For instance, 40-70% reductions in rotavirus diarrhea incidence reported in African settings after vaccine implementation [35] would translate to the prevention of approximately 12-20% of clinically-attended, antibiotic-treated diarrhea among children in GEMS. The recently-completed rotavirus Vaccine Impact on Diarrhea in Africa (VIDA) study [36], which assessed the incidence and etiology of MSD after rotavirus vaccine introduction at GEMS sites in The Gambia, Mali, and Kenya, will allow documentation of these effects.

Antibiotic use endpoints merit inclusion in trials of vaccines against enteric pathogens to demonstrate whether prevention of diarrhea meaningfully reduces overall antibiotic consumption [37]. Cluster-randomized trials and post-licensure studies may provide opportunities to assess whether the prevention of antibiotic use further reduces prevalence or burden of antimicrobial resistance in vaccine-targeted pathogens [38–40], and in commensal pathogens subjected to resistance selection [41]. Our findings help to define the burden of antibiotic use associated with enteropathogens for which vaccines are presently available or in the pipeline. These estimates should inform the prioritization of vaccines at stages spanning development, clinical evaluation, and implementation.

## Data Availability

Data from the GEMS1 and GEMS1A studies are available from https://clinepidb.org/ce/app/.

## ACKNOWLEDGMENTS

This work was funded by the Bill & Melinda Gates Foundation (grants OPP1190803 to RL and JAL, and OPP1033572 to KLK).

## SUPPORTING INFORMATION CAPTIONS

**S1 Text: Supporting information**. The supporting information document includes further elaborations of methods (sections 1-5), 10 supplemental tables, and one supplemental figure, and citations to references in the supplemental text, as listed in the Table of Contents (page 1).

